# The Effects of Adverse Life Events on Brain Development in the ABCD Study®: A Propensity-weighted Analysis

**DOI:** 10.1101/2024.09.25.24314355

**Authors:** Amanda Elton, Ben Lewis, Sara Jo Nixon

**Affiliations:** Department of Psychiatry, University of Florida, Gainesville, FL 32610, USA; Center for Addiction Research & Education, University of Florida, Gainesville, FL 32610, USA

## Abstract

Longitudinal studies of the effects of adversity on human brain development are complicated by the association of stressful events with confounding variables. To counter this bias, we apply a propensity-weighted analysis of the first two years of The Adolescent Brain Cognitive Development^SM^ (ABCD) Study® data, employing a machine learning analysis weighted by individuals’ propensity to experience adversity. Data included 338 resting-state functional connections from 7190 youth (46% female), divided into a training group (80%) and an independent testing group (20%). Propensity scores were computed using 390 variables to balance across two-year adverse life event exposures. Using elastic net regularization with and without inverse propensity weighting, we developed linear models in which changes in functional connectivity of brain connections during the two-year period served as predictors of the number of adverse events experienced during that same period. Haufe’s method was applied to forward-transform the backward prediction models. We also tested whether brain changes associated with adverse events correlated with concomitant changes in internalizing or externalizing behaviors or to academic achievement. In the propensity-weighted analysis, brain development significantly predicted the number of adverse events experienced during that period in both the training group (ρ=0.14, p<0.001) and the independent testing group (ρ=0.10, p<0.001). The predictor indicated a general pattern of decreased functional connectivity between large-scale networks and subcortical brain regions, particularly for cingulo-opercular and sensorimotor networks. These network-to-subcortical functional connectivity decreases inversely associated with the development of internalizing symptoms, suggesting adverse events promoted adaptive brain changes that may buffer against stress-related psychopathology. However, these same functional connections were also associated with poorer grades at the two-year follow-up. Although cortical-subcortical brain developmental responses to adversity potentially shield against stress-induced mood and anxiety disorders, they may be detrimental to other domains such as academic success.

## INTRODUCTION

Early life stress (ELS) increases risk for negative physical and mental health outcomes, such that a greater number of stressors experienced is associated with poorer outcomes in a dose- depended manner^1–4^. Stressful experiences in childhood can range from ongoing exposure to abuse, neglect, household dysfunction, discrimination, as well as discrete adverse and traumatic events. Such experiences are associated with alterations in brain structure and function, providing a mechanistic link to the development of stress-related psychopathologies, including depression, anxiety, and substance use disorders^5–9^. ELS-associated brain differences are largely recognized as representing perturbations in typical brain development leading to persistent alterations to brain structure and function^10,11^.

Human neuroimaging studies of brain development during childhood and adolescence have provided important insights into the dynamic changes in brain connectivity underlying typical development. For example, there is a significant increase in white matter connectivity between the cortical and subcortical brain regions during childhood and adolescence that supports cognitive and motor abilities, with prefrontal tracks implicated in executive control maturing in adolescence and adulthood^12–14^. Similarly, functional connectivity of the dorsal striatum with sensorimotor regions is present during childhood but diminishes through adolescence in favor of strengthening prefrontal-striatal connectivity^15^. Furthermore, increased resting-state functional connectivity between the prefrontal cortex and the amygdala during childhood and adolescence supports emotional regulation and reduces negative affect^16,17^. However, as children enter adolescence, the normative functional connectivity between the prefrontal cortex and amygdala switches from positive to negative, which is proposed to reflect maturing emotion regulation^18^.

Overall, studies suggest that changes in connectivity between cortical and subcortical brain regions during different periods of development is crucial for the development of motor, cognitive, and emotional processing abilities. Factors such as ELS may produce variations in the developmental trajectories of these processes, leading to risk or resilience for various negative health outcomes.

The ABCD Study® offers an unprecedented opportunity to prospectively investigate the effects of ELS on longitudinal brain development in a large and diverse sample of adolescents. For example, using ABCD data, Rakesh and colleagues examined effects of socioeconomic status on baseline resting-state connectivity of large-scale networks^19^. In that study, within and between-network connections involving sensorimotor networks and auditory network were particularly affected, in addition to multiple connections involving several frontal cognitive networks. However, because that study only included a single timepoint, prospective changes in brain development for these networks were not characterized. Another study by Brieant and colleagues using ABCD data examined developmental changes in functional connectivity associated with cumulative adverse life events experienced across the youth’s lifetime^20^. That analysis was focused on cingulo-opercular connectivity with the hippocampus and amygdala and identified reduced functional connectivity development in those connections. However, large-scale prospective studies of the impact of ELS on brain developmental changes have thus far been limited.

A major challenge to studying the effects of ELS through observational studies in humans is that exposure to stressors is typically associated with a multitude of other confounding variables that occur over a broad time period. For example, individuals exposed to one type of trauma are more likely to have also been previously exposed to other forms of stressors. The likelihood of experiencing stressors also varies by socioeconomic status, racial identity, family structure, and community environment. Furthermore, adverse childhood experiences related to living with family members with mental health disorders can be confounded by personal genetic risk for those disorders. These complex relationships have the potential to obscure and confound effects of stressful experiences on brain development. Although longitudinal designs can overcome some limitations, baseline brain measures may also systematically differ between those who are later exposed to stressors versus those who are not. To account for these myriad influences, propensity score-weighted analysis can adjust for potential confounding factors that may affect the accurate assessment of the relationship between adverse life events and brain development. Propensity score methods enable observational studies to provide effect estimates that more closely reflect those that would be obtained with randomized experimental designs^21^. Specifically, weighting individuals according to their propensity to experience adverse life events balances the distribution of covariates across the levels of this exposure (the “treatment”), thereby reducing the bias in effect estimates.

For the current study, we conducted a data-driven, propensity score-weighted analysis of ABCD data to examine the relationship between the number of adverse life events experienced during a two-year period and brain development during that same time period. Here we examine the effects of adverse life events on network-level brain functional connectivity derived from resting-state fMRI. In this study, we focused on acute stressors in the form of discrete adverse life events that occurred during the study timeframe (rather than lifetime events or chronic stress like low socioeconomic status), which allowed us to simulate, using propensity scores, an experimental design in which subjects are matched at baseline and prospectively randomized to an exposure. As a result, this design allowed us to more clearly attribute brain developmental effects over the study period to the stressful events experienced during that time period. This distinction in the timing of events is crucial since ELS exposures can have opposite effects depending on their developmental timing^7,22^. Using a machine learning approach, we identified patterns of functional connectivity development within and between cortical brain networks and subcortical regions that were associated with the number of adverse life events experienced between the baseline and two-year follow-up fMRI scans. Furthermore, we tested whether variations in stress-sensitive functional connections related to changes in behavior.

## METHODS

The present study utilized resting-state functional connectivity data from the ABCD study to investigate the impact of exposure to stressful events on brain functional development. For the current analysis, we downloaded data from data release 4.0 and included subjects who had complete resting-state fMRI data at both baseline and the two-year follow-up, providing a total sample size of 7,190 subjects. Subjects were an average 9.9±0.1 years old at baseline and 12.0±0.1 years old at follow-up.

Data were divided into a training group (80%; n=5,749) and a testing group (20%, n=1,441), with the site composition and adverse life events within each group constrained to be approximately equal. Using only the training data, we developed a resting-state functional connectivity-based classifier of adverse events that was subsequently evaluated on the independent testing group, as described in further detail below.

This analysis was not pre-registered. However, this was a data-driven analysis that did not test *a priori* hypotheses, and the inclusion of an independent testing group offers an important degree of rigor that demonstrates the robustness of the findings.

### Resting-state functional connectivity

Resting-state functional connectivity data were collected at baseline and a two-year follow-up. Data were downloaded as tabulated text files containing the pairwise Fisher z-transformed Pearson correlation coefficient denoting the functional connectivity between pairs of brain regions. Details of the processing of these data has been reported elsewhere^24^. Two sets of functional connectivity data were used: 1) the functional connectivity within and between 13 large-scale cortical brain networks (abcd_betnet02.txt) defined from the Gordon network parcellation^25^; and 2) the functional connectivity between the networks and 19 anatomically-defined subcortical regions (mrirscor02.txt). In total, 338 connections were considered.

To measure brain development over the two years between the baseline and follow-up scan, we calculated the residualized change in functional connectivity. First, functional connectivity estimates were cleaned to remove effects of variables known to systematically affect the functional connectivity estimates: For each time point independently, we used linear regression to estimate and then partial out effects of age in months at the time of the scan, the scanner manufacturer [Siemens (1) versus GE or Philips (0)], and the maximum and average framewise displacement and their squares (to remove both linear and nonlinear effects of motion). The adjusted functional connectivity values for the follow-up scan were then regressed on the corresponding adjusted values of the baseline scan, and the residuals (with the intercept added back) were retained. These calculations were conducted separately for the training and testing groups to avoid introducing bias into the test results.

### Adverse life events

We aimed to evaluate the effects of acute stress associated with specific adverse life events. Adverse life events were self-reported by participants at the 1-year follow-up and the 2-year follow-up on the Adverse Life Events Scale^26^. Participants indicted whether each of 25 events had ever occurred, had occurred within the past year, the perceived valence (“Was this a good or bad experience?”), and the perceived severity (“How much did the event affect you?”). Due to the known dose-effect relationship between ELS and psychiatric outcomes^1^, we focused our investigation on the total number of adverse life events. The negatively-rated events endorsed at the 1-year follow-up that had not occurred during the past year were counted as prior adverse life events. The total number of adverse life events experienced between the baseline scan and the 1-year follow-up was calculated as the total number of endorsed events rated as negative and occurring in the past year, regardless of the severity. This approach was similarly applied to estimate total adverse events between the 1- and 2-year follow-ups. The sum of adverse life events occurring between the baseline and 2-year follow-up was used in subsequent analyses.

### Propensity Score Calculation

Incorporating propensity scores into analyses offers a way to balance covariates across all levels of a variable of interest in an observational study as a way to approximate an experimental study in which subjects are randomly assigned to a “treatment.” This is an improved method compared with including many covariates in statistical models^27^. Propensity scores are first constructed in a statistical model to derive the predicted probabilities of treatment assignment based on the associated covariates. In this case, the “treatment” is the number of adverse life events. It is recommended to include covariates that predict not only the treatment but also the outcome (i.e., brain functional connectivity development) when constructing propensity scores^28^. Only baseline variables collected prior to the “treatment” are included. Following the calculation of propensity scores, one of several methods for incorporating propensity scores into the analysis can be implemented.

Here, propensity scores for the training group were calculated with the *weightit()* function in *R* statistical software using the propensity score method. There were 390 variables used to construct the propensity scores: study site, scanner manufacturer, baseline age, sex, race, ethnicity, number of prior adverse life events, T-scores for the internalizing and externalizing subscales of the Child Behavior Checklist (CBCL^29^) at baseline, puberty score at baseline, maternal and paternal histories of depression, maternal and paternal histories of alcohol and other substance use problems, parental education, income, whether children split their time between separate homes, baseline values for all included functional connections, and baseline in-scanner motion (mean and maximum framewise displacement and their squares). The relative contribution of each of these variables to the calculated propensity scores depends on the strength of their association with the number of adverse events. We used *weightit()* to calculate weights to balance confounding variables across the number of adverse life events, modeled as a Poisson distribution with a log link function. Here we use the inverse probability of treatment weighting method^30,31^, in which subjects for which the baseline variables were better predictors of the number of adverse events experienced were downweighted relative to subjects for which those variables were poorer predictors. As a result, we were able to reduce the confounding influence of these variables in our analysis. This weighting strategy was selected for its compatibility with continuous treatments.

### Elastic Net Prediction Analysis

We used an elastic net-regularized^32^ negative binomial regression to predict adverse life events using resting-state functional connectivity data with *glmregNB()* in *R* version 4.2.1. The prediction analysis was run twice: once inserting the propensity score weights in the *weights* option and once without weights to enable qualitative comparison. These analyses used 10-fold cross validation in which 90% of the training group was used to develop a functional connectivity predictor of stressful events for the remaining 10% of the training group, repeated 10 times.

Within each fold, a nested cross-validation tuning step involved selection of α and λ parameters: A series of 10 α values between 0.1 and 1 (in increments of 0.1) were tested to weight the penalty towards ridge or lasso optimization, where 1 is equivalent to lasso regularization.

Additionally, a series of 10 values for the regularization parameter, λ, was tested at each value of α, and another nested level of 10-fold cross-validation was used to select the values α and λ that provided the lowest cross-validation error. After fitting the set of selected functional connections to the 90% training subgroup, the linear equation was used to predict the number of adverse events experienced by the remaining 10%. This cross-validation procedure was repeated 10 times until all subjects in the training group had a predicted value for the number of adverse events experienced. These predicted values were then correlated with subjects’ actual values using a Spearman correlation analysis to evaluate the accuracy of the prediction. The 10-fold cross validation was repeated 5 times to ensure stability of the results across different partitions of the training data.

To combine the linear equations produced by each training iteration into the final prediction equation, we averaged coefficients across the iterations. Each subject’s data was entered into the final equation to obtain a predicted value. This final prediction equation was tested on both the training group (resubstitution, a biased estimate of accuracy) as well as the testing group (an unbiased estimate of accuracy) based on Spearman correlations between actual and predicted values.

To transform the backward prediction model (functional connectivity predicting adversity) to a forward model (adversity predicting functional connectivity), we used Haufe’s method for transforming multivariate neuroimaging results^33^. Because we only had one outcome variable in our model, this method involves calculating the covariance between each predictor and the predicted values, which produces transformed coefficients representing effects of adversity on functional connectivity development. Initially, we calculated the significance of these results by examining the magnitude of this transformed coefficient relative to both the variance within each 10-fold cross validation and between the 5 iterations to derive a t-statistic. There was a remarkable consistency across each cross validation and iteration, which resulted in 88% and 90% of the connections being deemed significant for the unweighted and propensity-weighted analyses, respectively (Supplemental Table 1). To improve interpretability of these findings, we adopt the recommendation to set effect sizes thresholds to focus on not only significant, but potentially important results^34^. Here we set Pearson’s r=0.50, which is generally considered a moderate effect size relationship^35^, as the minimum important effect size of the Haufe-transformed coefficients.

### Brain-Behavior Correlations

We next tested whether changes to brain connections associated with adversity may relate to changes in two functional domains: mental health and academic achievement.

To examine the implications of functional brain changes for mental health, we tested the association of significant functional connectivity predictors with parental reports of child behaviors using the Child Behavior Checklist (CBCL^29^) at the two-year follow-up. Specifically, we used T-scores for the internalizing and externalizing subscales of CBCL. Pearson correlations were calculated in separate analyses for the two subscales, partialing the corresponding baseline CBCL scores so that the correlations account for the relative change in behaviors over two years. We also partialed for the total number of adverse life events over the past two years in order to ensure that relationships between the brain and internalizing/externalizing behaviors were not driven by their common association with adverse life events.

To examine the implications of functional brain changes for academic achievement, we examined academic performance at the two-year follow-up. For this analysis, we used parent-reported grades, which were available as binned data, where greater values reflected poorer grades, and analyzed these relationships with Spearman correlations. Baseline scores were not available to enable a measure of change in academic performance. However, we partialed for the total number of adverse life events over the past two years.

### Sensitivity and Secondary Analyses

Motion during fMRI can affect functional connectivity estimates but is also closely associated with development and behavioral measures. We re-ran the elastic net prediction analysis including only subjects with lower levels of motion, which we defined as an average framewise displacement of <0.3 mm for both the baseline and follow-up scans. After applying this threshold, 3625 (63%) subjects remained in the training group and 893 (62%) remained in the testing group.

We next tested whether the effects of adverse life events on the brain substantially differed between males and females, as sex differences in the response to stress are well documented^36,37^. To test this possibility, we developed a predictor based only on females and predicted the number of adverse life events in males and vice versa. We used the same training and testing groups as the original analysis.

## RESULTS

### Elastic net prediction analysis

In the unweighted analysis, not correcting for propensity to experience adverse life events, the final prediction equation predicted the number of adverse life events in the training group (r=0.16, p<0.001) and the independent testing group (r=0.12, p<0.001). In the propensity-weighted analysis, the final prediction equation predicted the number of adverse life events in the training group (r=0.14, p<0.001) and the independent testing group (r=0.085, p=0.001). Table 2 displays the significant prediction accuracy results of the five iterations of the prediction analysis for both the training group and the independent testing group.

**Table 1.**
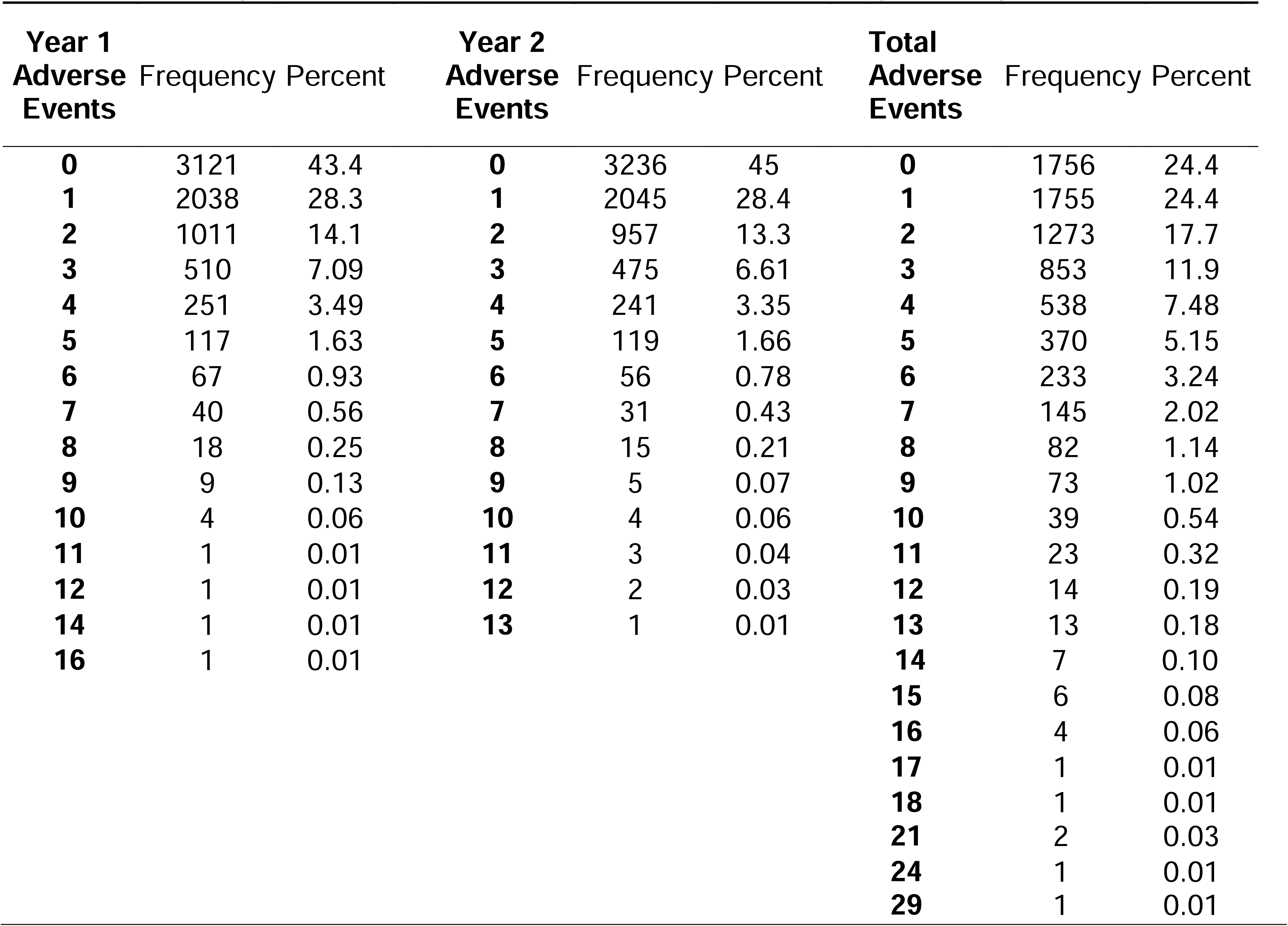
Frequency of adverse life events experienced over the two-year study period.

**Table 2.** Prediction accuracy across five iterations based on Spearman correlation between actual and predicted number of adverse life events.

| Unweighted | $\rho$ | 95% CI | | p | $\rho$ | 95% CI | | p |
| --- | --- | --- | --- | --- | --- | --- | --- | --- |
|  | Cross-Validation |  |  |  | Test group |  |  |  |
|  | 0.09 | 0.07 | 0.12 | <0.001 | 0.11 | 0.07 | 0.15 | <0.001 |
|  | 0.09 | 0.07 | 0.12 | <0.001 | 0.11 | 0.07 | 0.15 | <0.001 |
|  | 0.09 | 0.07 | 0.11 | <0.001 | 0.11 | 0.06 | 0.15 | <0.001 |
|  | 0.10 | 0.08 | 0.12 | <0.001 | 0.11 | 0.07 | 0.15 | <0.001 |
|  | 0.10 | 0.08 | 0.12 | <0.001 | 0.11 | 0.07 | 0.15 | <0.001 |
|  | Final |  |  |  |  |  |  |  |
|  | 0.15 | 0.13 | 0.18 | <0.001 | 0.11 | 0.07 | 0.15 | <0.001 |
| Propensity Weighted | $\rho$ | 95% CI | | p | $\rho$ | 95% CI | | p |
|  | Cross-Validation |  |  |  | Test group |  |  |  |
|  | 0.09 | 0.06 | 0.11 | <0.001 | 0.10 | 0.05 | 0.14 | <0.001 |
|  | 0.09 | 0.07 | 0.11 | <0.001 | 0.10 | 0.05 | 0.14 | <0.001 |
|  | 0.07 | 0.05 | 0.09 | <0.001 | 0.09 | 0.05 | 0.13 | <0.001 |
|  | 0.08 | 0.06 | 0.10 | <0.001 | 0.10 | 0.05 | 0.14 | <0.001 |
|  | 0.08 | 0.06 | 0.10 | <0.001 | 0.09 | 0.05 | 0.14 | <0.001 |
|  | Final |  |  |  |  |  |  |  |
|  | 0.14 | 0.12 | 0.16 | <0.001 | 0.10 | 0.05 | 0.14 | <0.001 |

In the unweighted analysis, 7 functional connections showed significant negative effects of adverse life events on development. For the propensity-weighted analysis, 10 functional connections showed negative effects, which included the 7 connections from the unweighted analysis in addition to three others (Figure 1). No connections showed significant positive effects in either analysis. Results are thresholded at |r|>0.50, which is intended to display only those connections with a moderate-to-large effect size relationship with the predictor. Supplemental Figure 1 displays the same data using an alternative, lower threshold (|r|>0.40). These results suggested an overall pattern in which the largest effects, which tended to be negative, were primarily observed between large scale brain networks and subcortical brain regions.

**Figure 1.**
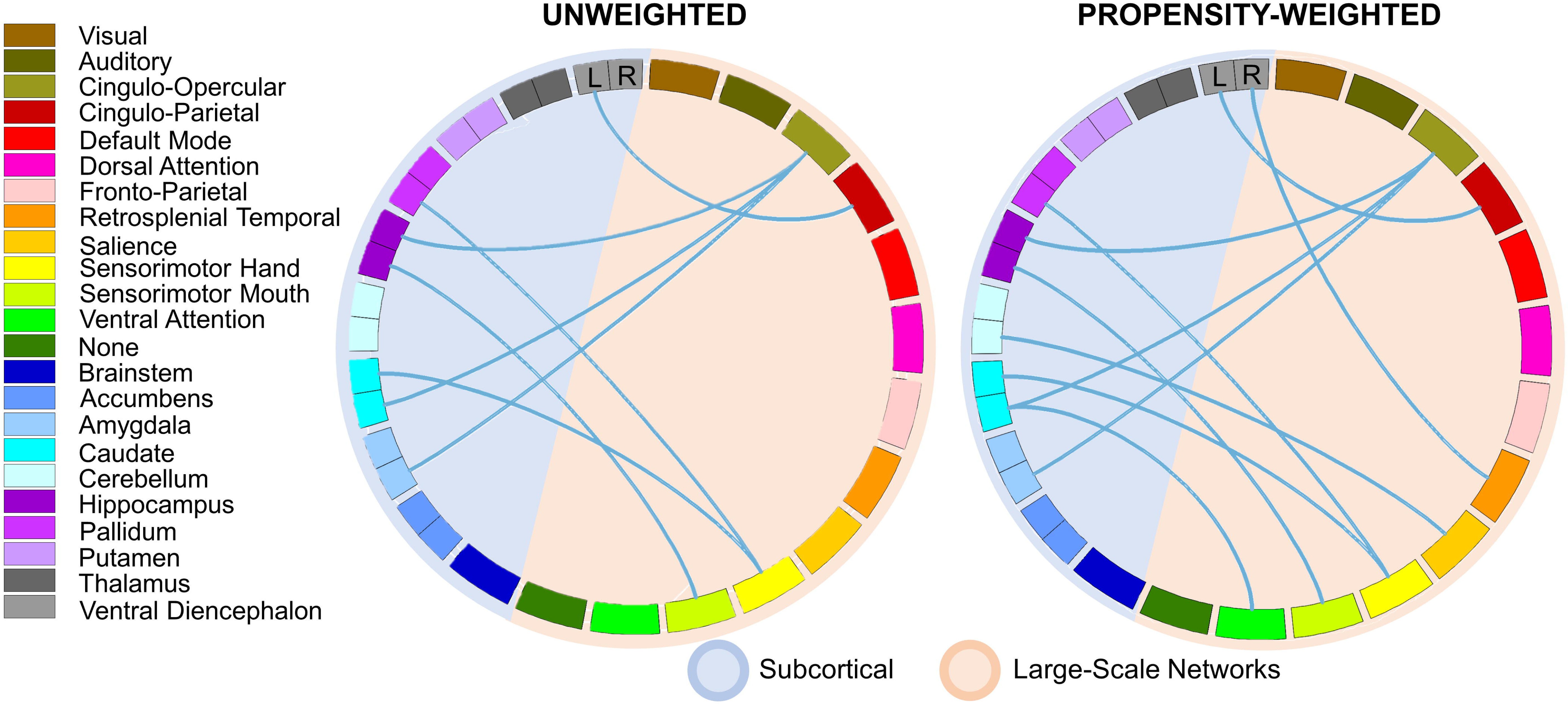
Circular plot of functional connections with significant negative (blue lines) associations with predicted values for the unweighted (left) and propensity weighted (right) analyses. There were no significant effects in the positive direction. All Plotted lines meet the threshold of |r|≥0.50 for the association between the connection and the predictor. Bilateral subcortical regions are plotted separately for left (L) and right (R) lateralization.

Figure 2 plots the baseline and follow-up values for each of the 10 significant functional connections from the propensity-weighted analysis. For visualization purposes, we perform a median split of the data, plotting subjects with <2 adverse life events and >2 adverse life events separately. Figure 2 also includes the residualized change values for each subgroup of the median split, which were calculated as the follow-up values after adjusting for the effect of baseline functional connectivity values estimated with linear regression. These connections are plotted using unweighted (Figure 2A) and propensity-weighted (Figure 2B) means. For each connection, there was a trend for functional connectivity to decrease between the baseline and two-year follow-up, regardless of stress exposure. However, this developmental decrease was accelerated in subjects reporting a greater number of adverse life events.

**Figure 2.**
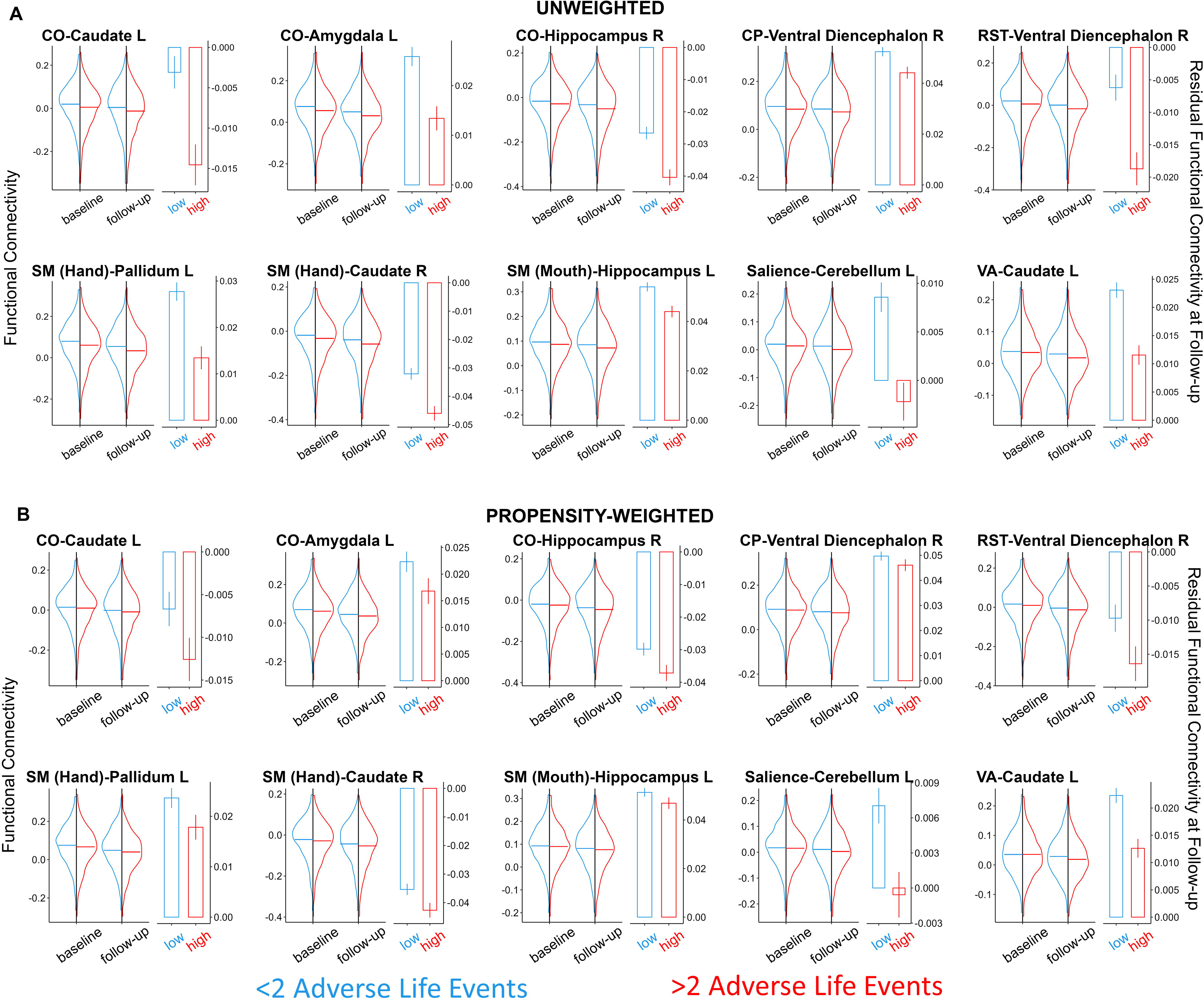
Functional connectivity by time point and number of adverse life events. Violin plots (left side of each panel) display the distributions and unweighted (A) or propensity-weighted (B) means of functional connectivity values at baseline and the two-year follow up for the pairwise connections significantly (|r|≥0.50) correlated with the predicted number of adverse life events based on the propensity-weighted analysis. Residualized change scores, which are functional connectivity values at the follow-up visit after adjusting via regression for functional connectivity at the baseline visit, are plotted to the right of each panel. Data are displayed for individuals with fewer (<2; blue) or greater (>2; red) negative life events during the two-year time period between scans based on a median split of the training sample. All displayed connections indicate a negative effect of adverse life events on development of functional connectivity. CO, cingulo-opercular; CP, cingulo-parietal; RST, retrosplenial-temporal; SM, sensorimotor; VA, ventral attention; L, left; R, right.

To interrogate whether certain networks were broadly affected by adverse life events, we calculated the average values for within-network connections (Figure 3A), between-network connections (Figure 3B), and network-to-subcortical connections (Figure 3C) for each network separately. These calculations included all connections for a given network, regardless of whether they met the threshold for significance. We also performed the same calculations for each subcortical brain region to all networks to examine whether there were subcortical regions that were particularly related to adverse life events (Figure 3D). Overall, adverse life events were related to reduced network-to-subcortical functional connectivity development across most networks, but particularly for the cingulo-opercular network and sensorimotor network. In addition to ELS effects on reduced network-to-subcortical connectivity, there was also a tendency for increased connectivity within and between motor and sensory networks. The patterns of covariance between all functional connections and the predictor for the propensity-weighted analysis was highly similar to that of the unweighted analysis, with a Pearson correlation across all connections yielding r=0.99. However, covariance values of the effects of ELS on these brain regions were generally reduced for the propensity-weighted analysis, as evidenced in Figure 3.

**Figure 3.**
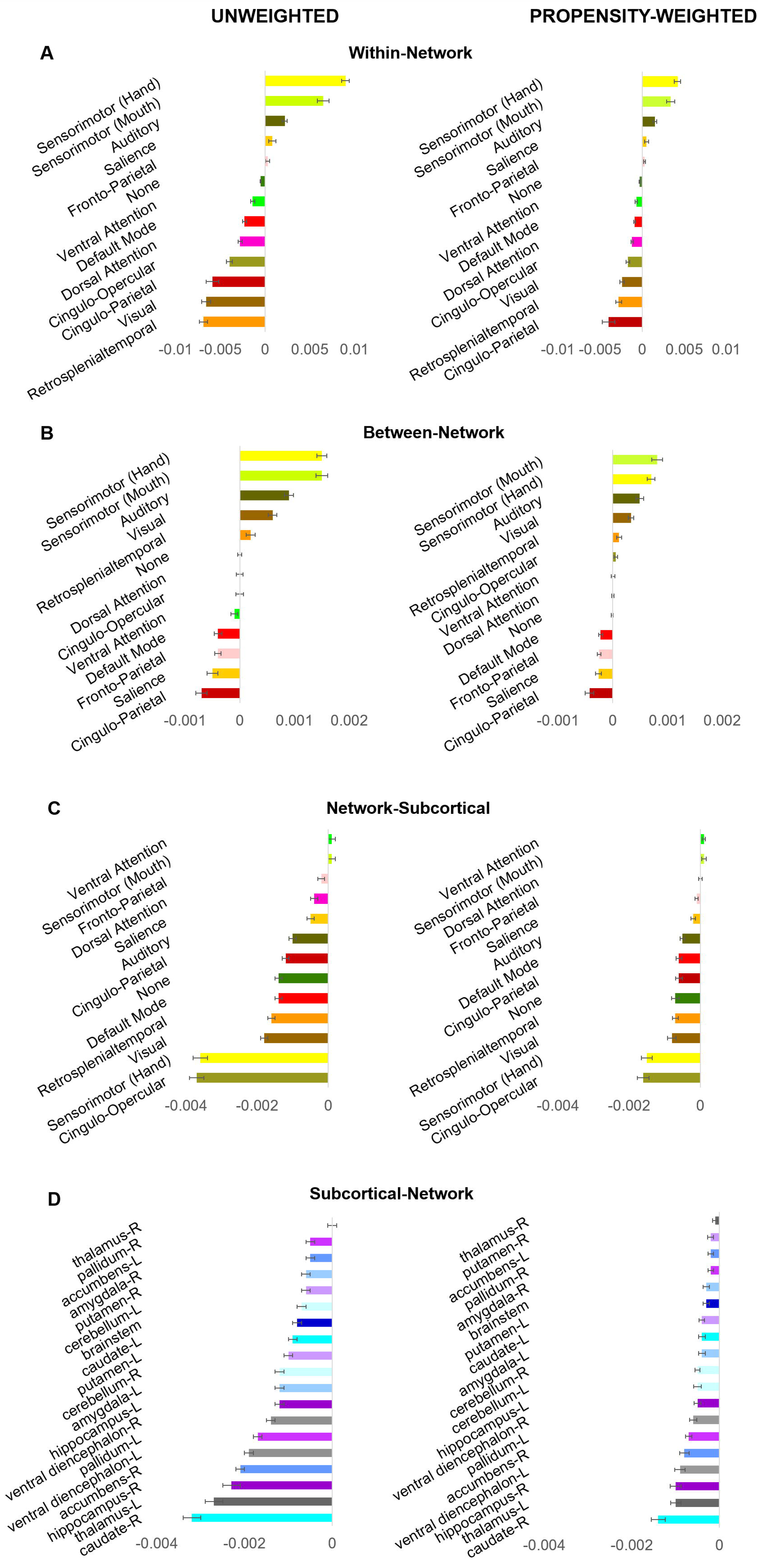
Estimated effects of adverse life events averaged by large-scale network for A) within-network functional connectivity, B) between-network functional connectivity, and C) network-to-subcortical functional connectivity. D) Estimated effects of adverse life events averaged by subcortical brain region for subcortical-to-network functional connectivity. Values are covariances between functional connectivity development and the predicted number of adverse life events, averaged across 5 iterations of 10-fold cross-validation.

### Brain-Behavior Correlations

A Pearson correlation between the number of adverse life events and CBCL internalizing and externalizing symptoms indicated that acute stressors were related to increased expression of both internalizing (r=0.054, p<0.001) and externalizing (r=0.082, p<0.001) behaviors, relative to baseline.

The correlations between brain functional connectivity development and changes in CBCL scores are presented in Table 3. Of the 10 functional connections tested, 6 demonstrated significant correlations with internalizing behaviors after FDR correction, whereas there were no significant effects for externalizing behaviors. However, these correlations were each in the positive direction. Among connections for which adverse life events were associated with lesser functional connectivity over time, decreases in connectivity were associated with reductions in internalizing symptoms. Thus, functional connectivity developmental changes associated with recent experiences of adverse life events at least partially protected against the development of internalizing psychopathology.

**Table 3.**
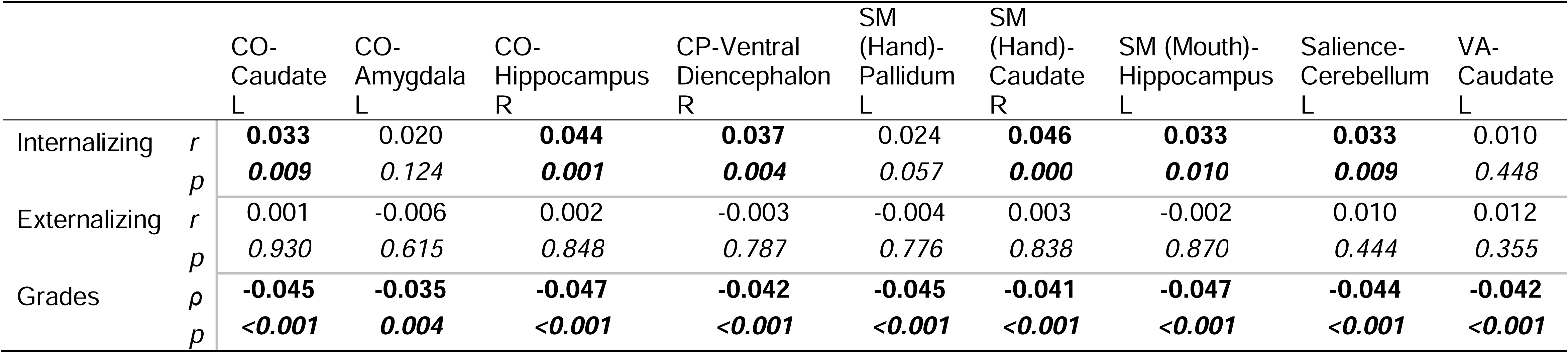
Correlations between brain functional connectivity changes and behavior.

A Spearman correlation between the number of adverse life events and parent-reported grades indicated that acute stressors were related to lower academic achievement (r=0.15, p<0.001). The correlations between brain functional connectivity development and grades is presented in Table 3. All 10 of the tested connections were significant, where functional connectivity developmental changes associated with recent experiences of adverse life events were associated with poorer grades.

### Sensitivity and Secondary Analyses

The elastic net prediction analysis of the low motion sample resulted in significant predictions, similar in strength to the full sample analysis (Supplemental Table 2). The pattern of covariance associations between the full set of connections and the predictor aligned closely with the results from the full sample (r=0.95, p<0.001, Supplemental Table 1), although the only significant functional connections using the |r|≥0.5 threshold extended from the sensorimotor (mouth) network to right putamen and salience network to left caudate.

The predictor developed using data from females was significantly correlated with the predictor developed on the males (r=0.86, p<0.001), although the significant connections were not fully consistent across the sexes (Supplemental Table 1). These sex-specific predictors were generally able to predict the number of adverse events across both males and females (Supplemental Table 3). Although the results do not preclude the possibility of significant sex differences, effects in females and males in this sample were largely similar.

## DISCUSSION

Using a data-driven approach with propensity score weighting, the current study indicated that adverse events in late childhood/early adolescence related to developmental reductions in cortical-subcortical functional connectivity. The findings primarily implicated connections between cortical networks and subcortical brain regions, more so than within or between networks. The affected cortical-subcortical connections include numerous subcortical structures extending beyond expected effects in the amygdala and hippocampus. Correlations with behavior suggested that the identified brain changes function potentially as adaptive mechanisms to counter stress-related development of mood and anxiety disorders, but may also be maladaptive for other domains such as academic achievement.

There were strong qualitative similarities between the propensity-weighted and unweighted analyses based on correlational analyses between the predictors. Although the implicated functional connections demonstrated robust effects of acute stress in both the unweighted and weighted analyses, the magnitude of effects were diminished after accounting for confounders. An inference of these weighting effects is that the variables that are closely associated with the propensity to experience acute stressors have qualitatively similar brain effects as the acute stressors themselves. Thus, analyses that do not adjust for these other variables are likely to over-estimate the effects of acute stressors for many brain connections, even though the estimated effects may be qualitatively accurate. Adjusting for propensity scores in studies of ELS can offer a method to more accurately quantify the impacts of various forms of stressors.

### Acceleration of maturation and developmental timing effects

A major finding of this study was that adverse life events related to decreases in functional connectivity between large-scale networks and subcortical regions. Similar decreases have been reported previously for exposure to chronic and lifetime stressors and have been described as an enhanced maturation of the brain^20,38,39^. Indeed, the direction of the effects in this study (Figure 2) demonstrates a general age-related reduction in functional connectivity among ELS-associated brain connections, with stronger effects in individuals exposed to a greater number of adverse life events. Notably, the current study extends previous reports by revealing that acute stressors also promote these maturational effects, as these effects persisted when adjusting for chronic stressors and other variables associated with the propensity to experience adverse life events, such as total lifetime adverse events and socioeconomic status.

Nonetheless, the observed effects may be specific to the developmental period included in this analysis, as different brain systems may be susceptible to ELS exposure at different ages^7^. For example, ELS related to neighborhood environment is associated with enhanced brain maturation (measured by a brain age index) during early adolescence (∼12 years) as seen in the current study, but these effects are no longer present by late adolescence (∼20 years)^39^.

Furthermore, in contrast with the current study, exposure to ELS earlier in childhood^40^ or later in adolescence^41^ and early adulthood^42^ has been associated with *increases* in cortical-subcortical functional connectivity. Thus, the developmental processes occurring during the time of stress exposure are likely to influence the observed effects.

For example, the developmental window for this analysis (9∼12 years) is a key time period when threat-regulation responses involving cortical-hippocampal-amygdala circuits are developing^43^. Indeed, connections between the cingulo-opercular network and hippocampus and amygdala were among the most affected. Although previous work in a smaller sample suggested that the maturational effects of ELS are specific to these threat and emotion regulation circuits^38^, the current findings suggest much broader effects. Specifically, there was a general trend for nearly all of the networks and subcortical regions to exhibit diminished network-to-subcortical connectivity in response to ELS (Figure 3C, 3D). In fact, the functional connectivity between sensorimotor networks and subcortical brain regions showed some of the strongest effects. Children exhibit stronger functional connections between primary sensory networks and subcortical brain regions relative to adults^44^. Therefore, the reductions in connectivity between sensorimotor regions and subcortical structures is consistent with an enhanced developmental trajectory. The effects of ELS on early-developing motor networks and threat regulation circuits is consistent with the notion that certain brain regions are particularly susceptible to experiences during specific “critical periods” of development^45,46^. Future work should examine whether brain functional connectivity changes observed in the current study in response to acute stressors lead to persistent effects in later adolescence, even in the absence of repeated exposures.

### Key role of the cingulo-opercular network

The network that demonstrated some of the greatest ELS-related developmental effects on subcortical functional connectivity was the cingulo-opercular network. This network is comprised on functional connections between the dorsal cingulate cortex, dorsal anterior insula/frontal operculum, and thalamus. It is generally considered a “task control” network, with a specific role in tonic alertness^47,48^. Of the subcortical regions tested, the amygdala and hippocampus demonstrated the strongest functional connectivity decreases with the cingulo-opercular network, similar to previously-reported effects of cumulative life events^20^. ELS has been repeatedly associated with reduced hippocampal volume^46,49^ and altered amygdala reactivity to emotional stimuli^22,50,51^, as well as altered connectivity between these regions and the prefrontal cortex^52^. Preclinical and human work suggests that these effects are at least partly related to effects of stress hormones^53^. Given the role of the cingulo-opercular network in maintaining alertness to the external environment, connectivity to these stress-sensitive subcortical regions may underly hypervigilance associated with the development of stress-related disorders.

### Relationship to symptoms and grades

Although adverse life events were associated with greater internalizing and externalizing psychopathology, paradoxically, the functional connectivity changes following such events were related to decreased internalizing psychopathology. These correlational analyses suggest that the brain developmental effects identified in this study may be an adaptive response to ELS exposure. These outcomes are reinforced by previous work showing that greater cingulo-opercular network functional connectivity was associated with heightened internalizing symptomatology in the ABCD study at baseline (functional connectivity with putamen)^54^ and at the 2-year follow-up (functional connectivity with bilateral amygdala and right hippocampus)^20^. By controlling for the number of adverse events experienced as well as baseline symptoms, the current analysis supports the notion that brain development can moderate the extent to which internalizing symptoms are expressed in response to adversity. Intriguingly, the neural correlates of the *increased* internalizing symptomatology in response to ELS were not identified. In addition, identified changes were conversely maladaptive in the context of the academic environment, as lower functional connectivity was associated with poorer grades. These findings highlight how brain changes that allow adolescents to adapt to stressors in their current environment may ultimately be maladaptive for other functional domains and could potentially be detrimental in the longterm^7^.

### Limitations

This study identified effects of discrete adverse events on brain development in a specific developmental window, which may not generalize to other developmental periods and other forms of stress. Furthermore, we did not differentiate between domains of stressful events in this study, despite some evidence that dimensions related to deprivation and threat may yield different brain effects^49,55^. Future work could also consider effects of perceived severity of stressors. Additionally, correlational effect sizes between resting-state connectivity and behavioral measures are typically small, as observed in the current analysis^23^. Neural changes in response to adversity may be inadequately captured by coarse measures like BOLD fMRI. However, the utility of identifying such correlations is not limited to characterizing how much variance fMRI signals account for in relationships between adversity and behavioral outcomes, but also in identifying patterns of brain effects (i.e., primarily cortical-to-subcortical) and directionality of behavioral correlations, which offer important clues as to how the brain responds to adversity. Finally, data collection for the ABCD study is ongoing, so the impact of the identified brain changes on future health and behaviors remains to be determined.

## Conclusions

Adverse events can have acute effects on brain function, leading to greater maturation of connections between cortical and subcortical brain regions. However, this more rapid development is associated with both costs and benefits. These findings update our understanding of the complex interplay between risk and resilience processes in the brain, and understanding these nuanced effects will be important for designing interventions to mitigate the impact of adversity on brain and behavioral outcomes.

## Data Availability

All data produced in the present study are available upon reasonable request to the authors

## ACKNOWLEDGEMENTS

Data used in the preparation of this article were obtained from the Adolescent Brain Cognitive Development^SM^ (ABCD) Study (https://abcdstudy.org), held in the NIMH Data Archive (NDA). This is a multisite, longitudinal study designed to recruit more than 10,000 children age 9-10 and follow them over 10 years into early adulthood. The ABCD Study is supported by the National Institutes of Health and additional federal partners under award numbers U01DA041048, U01DA050989, U01DA051016, U01DA041022, U01DA051018, U01DA051037, U01DA050987, U01DA041174, U01DA041106, U01DA041117, U01DA041028, U01DA041134, U01DA050988, U01DA051039, U01DA041156, U01DA041025, U01DA041120, U01DA051038, U01DA041148, U01DA041093, U01DA041089, U24DA041123, U24DA041147. A full list of supporters is available at https://abcdstudy.org/federal-partners.html. A listing of participating sites and a complete listing of the study investigators can be found at https://abcdstudy.org/consortium_members/. ABCD consortium investigators designed and implemented the study and/or provided data but did not necessarily participate in the analysis or writing of this report. This manuscript reflects the views of the authors and may not reflect the opinions or views of the NIH or ABCD consortium investigators. The ABCD data repository grows and changes over time. The ABCD data used in this report came from 10.15154/1523041. DOIs can be found at https://doi.org/10.15154/1523041. AE was supported by K01AA026334. BL was supported by K01AA026893.

## REFERENCES

1 Felitti, V. J. et al. Relationship of Childhood Abuse and Household Dysfunction to Many of the Leading Causes of Death in Adults: The Adverse Childhood Experiences (ACE) Study. American Journal of Preventive Medicine 14, 245–258 (1998). Doi: 10.1016/s0749-3797(98)00017-8

2 Dube, S. R. et al. Adverse childhood experiences and the association with ever using alcohol and initiating alcohol use during adolescence. Journal of Adolescent Health 38, 444.e441–444.e410 (2006).

3 Enoch, M.-A. The role of early life stress as a predictor for alcohol and drug dependence. Psychopharmacology 214, 17–31 (2011).

4 Anda, R. F. et al. Adverse childhood experiences, alcoholic parents, and later risk of alcoholism and depression. Psychiatr Serv 53, 1001–1009 (2002).

5 Pechtel, P. & Pizzagalli, D. A. Effects of Early Life Stress on Cognitive and Affective Function: An Integrated Review of Human Literature. Psychopharmacology (Berl) 214, 55–70 (2011). 10.1007/s00213-010-2009-2

6 McLaughlin, K. A. et al. Widespread reductions in cortical thickness following severe early-life deprivation: a neurodevelopmental pathway to attention-deficit/hyperactivity disorder. Biol Psychiatry 76, 629–638 (2014). 10.1016/j.biopsych.2013.08.016

7 Teicher, M. H., Samson, J. A., Anderson, C. M. & Ohashi, K. The effects of childhood maltreatment on brain structure, function and connectivity. Nature Reviews Neuroscience 17, 652–666 (2016). doi:10.1038/nrn.2016.111

8 Kaiser, R. H. et al. Childhood stress, grown-up brain networks: corticolimbic correlates of threat-related early life stress and adult stress response. Psychological Medicine 48, 1157–1166 (2018). 10.1017/S0033291717002628

9 Taylor, S. E. Mechanisms linking early life stress to adult health outcomes. Proceedings of the National Academy of Sciences of the United States of America 107, 8507–8512 (2010). 10.1073/pnas.1003890107

10 Malter Cohen, M., et al. Early-life stress has persistent effects on amygdala function and development in mice and humans. Proc Natl Acad Sci U S A 110, 18274–18278 (2013). 10.1073/pnas.1310163110

11 Boecker, R. et al. Impact of early life adversity on reward processing in young adults: EEG-fMRI results from a prospective study over 25 years. PloS one 9, e104185 (2014). 10.1371/journal.pone.0104185

12 Simmonds, D. J., Hallquist, M. N., Asato, M. & Luna, B. Developmental stages and sex differences of white matter and behavioral development through adolescence: A longitudinal diffusion tensor imaging (DTI) study. NeuroImage 92, 356–368 (2014). 10.1016/j.neuroimage.2013.12.044

13 Lebel, C., Walker, L., Leemans, A., Phillips, L. & Beaulieu, C. Microstructural maturation of the human brain from childhood to adulthood. NeuroImage 40, 1044–1055 (2008). 10.1016/j.neuroimage.2007.12.053

14 Asato, M. R., Terwilliger, R., Woo, J. & Luna, B. White Matter Development in Adolescence: A DTI Study. Cerebral Cortex 20, 2122–2131 (2010). 10.1093/cercor/bhp282

15 Choi, E. J., Vandewouw, M. M., de Villa, K., Inoue, T. & Taylor, M. J. The development of functional connectivity within the dorsal striatum from early childhood to adulthood. Developmental Cognitive Neuroscience 61 (2023). ARTN 101258 10.1016/j.dcn.2023.101258

16 Gaffrey, M. S., Barch, D. M., Luby, J. L. & Petersen, S. E. Amygdala Functional Connectivity Is Associated With Emotion Regulation and Amygdala Reactivity in 4-to 6-Year-Olds. Journal of the American Academy of Child and Adolescent Psychiatry 60, 176–185 (2021). 10.1016/j.jaac.2020.01.024

17 Connolly, C. G. et al. Resting-state functional connectivity of the amygdala and longitudinal changes in depression severity in adolescent depression. Journal of Affective Disorders 207, 86–94 (2017). 10.1016/j.jad.2016.09.026

18 Gee, D. G. et al. A Developmental Shift from Positive to Negative Connectivity in Human Amygdala-Prefrontal Circuitry. Journal of Neuroscience 33, 4584–4593 (2013). 10.1523/Jneurosci.3446-12.2013

19 Rakesh, D., Zalesky, A. & Whittle, S. Similar but distinct - Effects of different socioeconomic indicators on resting state functional connectivity: Findings from the Adolescent Brain Cognitive Development (ABCD) Study. Developmental Cognitive Neuroscience 51 (2021). ARTN 101005 10.1016/j.dcn.2021.101005

20 Brieant, A. E., Sisk, L. M. & Gee, D. G. Associations among negative life events, changes in cortico-limbic connectivity, and psychopathology in the ABCD Study. Developmental cognitive neuroscience 52, 101022 (2021). 10.1016/j.dcn.2021.101022

21 Rosenbaum, P. R. & Rubin, D. B. The Central Role of the Propensity Score in Observational Studies for Causal Effects. Biometrika 70, 41–55 (1983). DOI 10.1093/biomet/70.1.41

22 Zhu, J. J., Anderson, C. M., Ohashi, K., Khan, A. & Teicher, M. H. Potential sensitive period effects of maltreatment on amygdala, hippocampal and cortical response to threat. Molecular Psychiatry (2023). 10.1038/s41380-023-02002-5

23 Marek, S. et al. Reproducible brain-wide association studies require thousands of individuals (Mar, 10.1038/s41586-022-04492-9, 2022). Nature 605, E11-E11 (2022). 10.1038/s41586-022-04692-3

24 Hagler, D. J. et al. Image processing and analysis methods for the Adolescent Brain Cognitive Development Study. NeuroImage 202 (2019). ARTN 116091 10.1016/j.neuroimage.2019.116091

25 Gordon, E. M. et al. Generation and Evaluation of a Cortical Area Parcellation from Resting-State Correlations. Cerebral Cortex 26, 288–303 (2016). 10.1093/cercor/bhu239

26 Tiet, Q. Q. et al. Adverse life events and resilience. J Am Acad Child Adolesc Psychiatry 37, 1191–1200 (1998). 10.1097/00004583-199811000-00020

27 Elze, M. C. et al. Comparison of Propensity Score Methods and Covariate Adjustment Evaluation in 4 Cardiovascular Studies. J Am Coll Cardiol 69, 345–357 (2017). 10.1016/j.jacc.2016.10.060

28 Brookhart, M. A. et al. Variable selection for propensity score models. Am J Epidemiol 163, 1149–1156 (2006). 10.1093/aje/kwj149

29 Achenbach, T. M., Verhulst, F. C., Baron, G. D. & Althaus, M. A comparison of syndromes derived from the Child Behavior Checklist for American and Dutch boys aged 6-11 and 12-16. J Child Psychol Psychiatry 28, 437–453 (1987). 10.1111/j.1469-7610.1987.tb01765.x

30 Austin, P. C. An Introduction to Propensity Score Methods for Reducing the Effects of Confounding in Observational Studies. Multivariate Behavioral Research 46, 399–424 (2011). Pii 938470000 10.1080/00273171.2011.568786

31 Lunceford, J. K. & Davidian, M. Stratification and weighting via the propensity score in estimation of causal treatment effects: a comparative study. Stat Med 23, 2937–2960 (2004). 10.1002/Sim.1903

32 Zou, H. & Hastie, T. Regularization and variable selection via the elastic net. Journal of the Royal Statistical Society: Series B (Statistical Methodology) 67, 301–320 (2005). 10.1111/j.1467-9868.2005.00503.x

33 Haufe, S. et al. On the interpretation of weight vectors of linear models in multivariate neuroimaging. NeuroImage 87, 96–110 (2014). 10.1016/j.neuroimage.2013.10.067

34 Dick, A. S. et al. Meaningful associations in the adolescent brain cognitive development study. NeuroImage 239 (2021). ARTN 118262 10.1016/j.neuroimage.2021.118262

35 Cohen, J. A power primer. Psychol Bull 112, 155–159 (1992). 10.1037//0033-2909.112.1.155

36 Elton, A. et al. Childhood Maltreatment is Associated with a Sex-Dependent Functional Reorganization of a Brain Inhibitory Control Network. Human Brain Mapping 35, 1654–1667 (2014). 10.1002/hbm.22280

37 Kudielka, B. M. & Kirschbaum, C. Sex differences in HPA axis responses to stress: a review. Biological Psychology 69, 113–132 (2005). 10.1016/j.biopsycho.2004.11.009

38 Herzberg, M. P. et al. Accelerated maturation in functional connectivity following early life stress: Circuit specific or broadly distributed? Developmental cognitive neuroscience 48, 100922 (2021). 10.1016/j.dcn.2021.100922

39 Rakesh, D. et al. Neighborhood disadvantage and longitudinal brain-predicted-age trajectory during adolescence. Developmental cognitive neuroscience 51, 101002 (2021). 10.1016/j.dcn.2021.101002

40 Barch, D. et al. Effect of Hippocampal and Amygdala Connectivity on the Relationship Between Preschool Poverty and School-Age Depression. American Journal of Psychiatry 173, 625–634 (2016). 10.1176/appi.ajp.2015.15081014

41 Rakesh, D. et al. Unraveling the Consequences of Childhood Maltreatment: Deviations From Typical Functional Neurodevelopment Mediate the Relationship Between Maltreatment History and Depressive Symptoms. Biol Psychiat-Cogn N 6, 329–342 (2021). 10.1016/j.bpsc.2020.09.016

42 Ilomaki, M. et al. Early life stress is associated with the default mode and fronto-limbic network connectivity among young adults. Front Behav Neurosci 16 (2022). ARTN 958580 10.3389/fnbeh.2022.958580

43 Uhlhaas, P. J. et al. Towards a youth mental health paradigm: a perspective and roadmap. Mol Psychiatry (2023). 10.1038/s41380-023-02202-z

44 Supekar, K., Musen, M. & Menon, V. Development of Large-Scale Functional Brain Networks in Children. Plos Biology 7 (2009). ARTN e1000157 10.1371/journal.pbio.1000157

45 Larsen, B., Sydnor, V. J., Keller, A. S., Yeo, B. T. T. & Satterthwaite, T. D. A critical period plasticity framework for the sensorimotor-association axis of cortical neurodevelopment. Trends Neurosci (2023). 10.1016/j.tins.2023.07.007

46 Teicher, M. H. et al. Differential effects of childhood neglect and abuse during sensitive exposure periods on male and female hippocampus. NeuroImage 169, 443–452 (2018). 10.1016/j.neuroimage.2017.12.055

47 Sadaghiani, S. & D’Esposito, M. Functional Characterization of the Cingulo-Opercular Network in the Maintenance of Tonic Alertness. Cerebral Cortex 25, 2763–2773 (2015). 10.1093/cercor/bhu072

48 Coste, C. P. & Kleinschmidt, A. Cingulo-opercular network activity maintains alertness. NeuroImage 128, 264–272 (2016). 10.1016/j.neuroimage.2016.01.026

49 Machlin, L. et al. Distinct Associations of Deprivation and Threat With Alterations in Brain Structure in Early Childhood. Journal of the American Academy of Child and Adolescent Psychiatry 62, 885-+ (2023). 10.1016/j.jaac.2023.02.006

50 van Harmelen, A.-L. et al. Enhanced amygdala reactivity to emotional faces in adults reporting childhood emotional maltreatment. Social Cognitive and Affective Neuroscience (2012). 10.1093/scan/nss007

51 Williams, L. M. et al. Trauma modulates amygdala and medial prefrontal responses to consciously attended fear. NeuroImage 29, 347–357 (2006). 10.1016/j.neuroimage.2005.03.047

52 Savulich, G. et al. Effects of naltrexone are influenced by childhood adversity during negative emotional processing in addiction recovery. Transl Psychiatry 7, e1054- (2017). 10.1038/tp.2017.34

53 Veer, I. M. et al. Endogenous cortisol is associated with functional connectivity between the amygdala and medial prefrontal cortex. Psychoneuroendocrinology 37, 1039–1047 (2012). 10.1016/j.psyneuen.2011.12.001

54 Lees, B. et al. Altered Neurocognitive Functional Connectivity and Activation Patterns Underlie Psychopathology in Preadolescence. Biol Psychiat-Cogn N 6, 387–398 (2021). 10.1016/j.bpsc.2020.09.007

55 Sheridan, M. A. et al. Neural Structure Is Independently Predicted by Deprivation and Threat in Early Childhood. Journal of the American Academy of Child and Adolescent Psychiatry 58, S340–S340 (2019). 10.1016/j.jaac.2019.07.808

