## Supplementary figures and images for "The Effects of Adverse Life Events on Brain Development in the ABCD Study®: A Propensity-weighted Analysis"

### Supplemental Figure 1

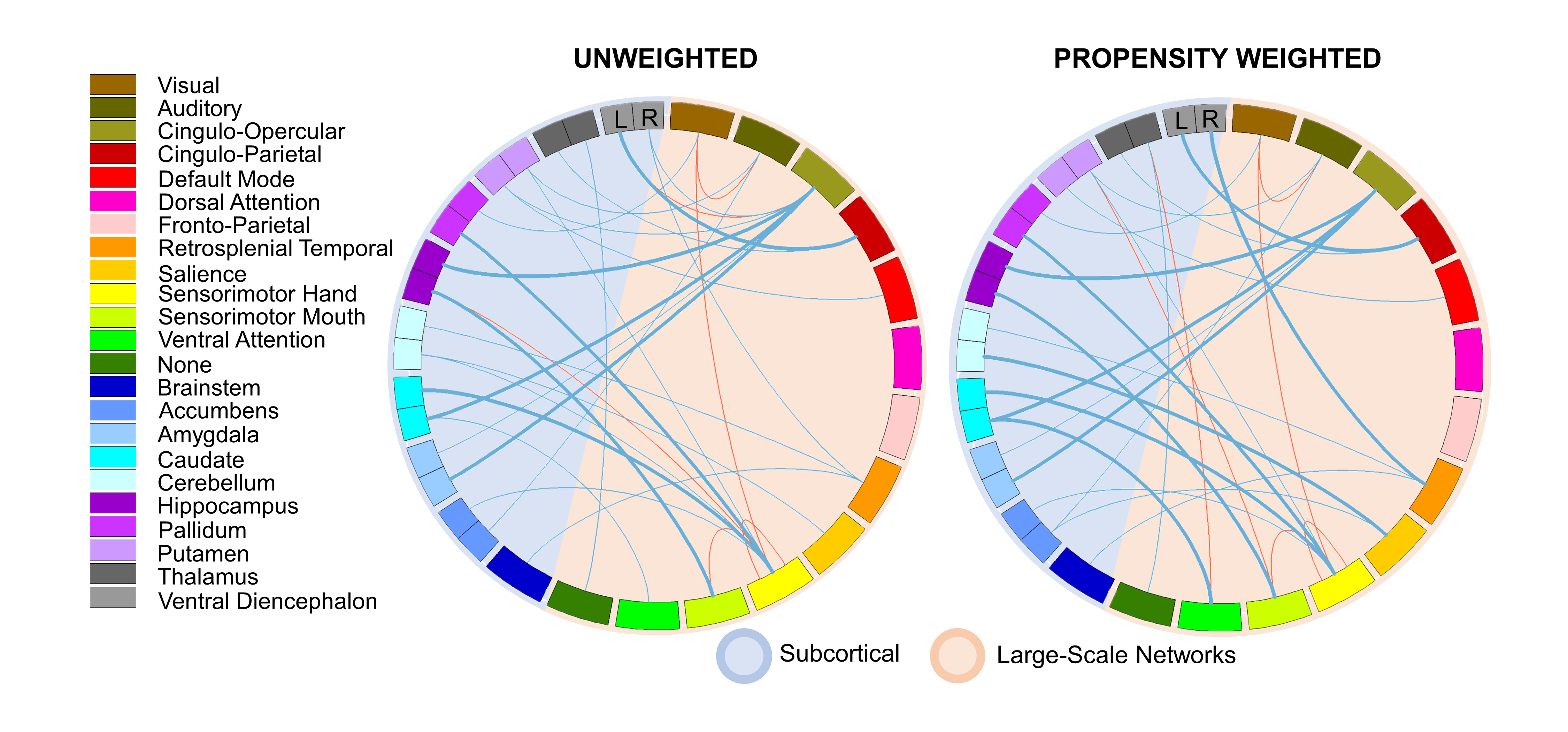
